# A novel Vascular Leak Index identifies sepsis patients with a higher risk for in-hospital death and fluid accumulation

**DOI:** 10.1101/2021.08.22.21262080

**Authors:** Jay Chandra, Miguel Ángel Armengol, Gwendolyn Lee, Alexandria Lee, Patrick Thoral, Paul Elbers, Hyung-Chul Lee, John S. Munger, Leo Anthony Celi, David A. Kaufman

## Abstract

**Purpose:** Sepsis is a leading cause of morbidity and mortality worldwide and is characterized by vascular leak syndrome. Treatment for sepsis, specifically intravenous fluids, may worsen deterioration in the context of vascular leak.

**Methods:** We performed a retrospective cohort study of sepsis patients in four ICU databases in North America, Europe, and Asia. We developed an intuitive vascular leak index (VLI) and determined the relationship between VLI and in-hospital death and 36h-84h fluid balance using generalized additive models (GAM).

**Results:** Using GAM, we found that increased VLI is associated with an increased risk of in-hospital death. Patients with a VLI in the highest quartile (Q4), across the four datasets, had a 1.61-2.31 times increased odds of dying in the hospital compared to patients with a VLI in the lowest quartile (Q1). VLI Q2 and Q3 were also associated with increased odds of dying. The relationship between VLI, treated as a continuous variable, and in-hospital death and 36h-84h death was statistically significant in the three datasets with a large number of patients. Specifically, we observed that as VLI increased, there was increase in the risk for in-hospital death and 36h-84h fluid balance. For the few patients with a positive VLI, this relationship differed across databases.

**Conclusions:** Our VLI identifies groups of patients who may be at higher risk for in-hospital death and for fluid accumulation early in the ICU course. This relationship persisted in models developed to control for severity of illness and chronic comorbidity burden.

## INTRODUCTION

Sepsis is a leading cause of morbidity and mortality worldwide. In 2017, there were approximately 49 million sepsis cases worldwide, resulting in 11 million related deaths [1]–[5]. Sepsis is also costly, accounting for over $24 billion in annual hospital costs in the United States. Costs are also rising: one study found an increase in $1.5 billion to treat patients with hospital-associated sepsis over the three-year period from 2015 to 2018 [6].

Expert guidelines recommend infusion of intravenous (IV) fluids to increase venous return, cardiac stroke volume, cardiac output, and ultimately tissue perfusion [7]. Clinical studies, however, suggest that fewer than half of hemodynamically unstable patients respond to fluids when fluid response is defined as an increase in the cardiac output of more than 10% [8]. Even in patients who are fluid responsive, IV fluids may be ineffective at improving tissue perfusion. Sepsis and other severe inflammatory states are often characterized by increased vascular permeability, termed the “vascular leak syndrome.” This results in physiologic derangements such as low circulating blood volume, impaired drug-binding capacity due to loss of plasma proteins, and organ failure due to tissue edema. Multiple studies associate a positive fluid balance with impaired organ function and a higher risk of death [9]–[11]. Expert guideline-recommended treatment of sepsis-induced hypotension includes administering IV fluids and vasopressors, either of which might be harmful to individual patients depending on the specific clinical situation [12], [13].

Measuring vascular leakage of fluid and protein requires special equipment that may not be widely available. Some investigators have proposed that the degree of vascular leak can be inferred by measuring hematocrit levels as IV fluids are infused. Hematocrit represents the concentration of hemoglobin, a protein too large to leak out of the vasculature. If infused fluid remains within the vasculature, hematocrit levels decline, as plasma volume increases and hemodilution occurs. If fluid leaks from the vasculature, the hematocrit should decline more slowly or even increase.

The goal of this investigation was to formulate an easily-calculated vascular leak index (VLI) that clinicians can use to estimate prognosis and guide care. We hypothesized that increasing VLI would be associated with a more positive fluid balance and an increased risk for death. We aimed to refine existing definitions of VLI by factoring in other key characteristics and clinical measurements [14], [15]. We derived and validated a VLI by evaluating the hematocrit over time with respect to the volume of IV fluids infused.

## METHODS

### Study Population

We conducted a retrospective analysis of patients from the eICU, MIMIC-III, AmsterdamUMCdb, and SNUH databases [16]–[20]. Details on the patient cohort contained in each dataset and the relevant Institutional Review Board information for the de-identified data is contained in the supplementary materials.

Our study population includes patients diagnosed with sepsis by ICD9 and Angus criteria in the eICU-CRD, MIMIC-III [21]. In AmsterdamUMCdb, diagnosis of sepsis at admission, diagnosis of other severe infections, the use of antibiotics not for prophylaxis after surgery, and finally the presence of sepsis cultures were all used to identify sepsis patients. In the SNUH database, due to data constraints, we only used diagnosis of sepsis at admission to identify sepsis patients. We excluded patients who were diagnosed with bleeding by ICD9, had other excess fluid output (if the data was available), were undergoing renal replacement therapy, or received blood products. In addition, patients who were missing VLI-related data including fluid balance, height, weight, and hematocrit, were excluded. In the eICU database, we only included patients in hospitals that we identified as having reliable fluid intake and output data.

### Vascular Leak Index

We developed an intuitive equation for VLI based on relevant variables, including hematocrit levels at two time points during ICU care and net volume of fluid administered. We reasoned that the relationship between the volume of fluid infused and the change in hematocrit would yield information about how much fluid remained in or escaped from the vascular space. Specifically, the change in the hematocrit divided by the net fluid balance would quantify vascular leak. To normalize for differences among patients’ blood volume, we divided the fluid volume by each patient’s body surface area, as suggested by Nadler and colleagues (Equation 1) [22]. We chose a multiplication factor of 1000 for easier interpretability. Only patients with positive fluid balance were included in the final patient cohort, because they have an increased risk of death and severity of illness [23]. Finally, given the difficulty in recording fluid intake and output, we conducted median imputation for the lower and upper 5% quantiles of VLI.

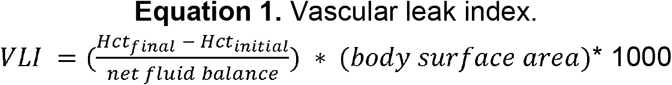

### Variables and Outcomes

Initial hematocrit values were measured at study baseline (0 h), including the range during the previous 12 h when the patient may have been admitted to the emergency department through the following 18 h. The first hematocrit during this timeframe was considered the initial value. Final hematocrit levels were calculated as the average among hematocrit values measured between 18 h and 36 h. The volume of total fluids administered between the previous 6 h prior to ICU admission and 36 h was recorded. Urine output was recorded and the net fluid balance was calculated as total fluid input minus urine output.

Patient outcomes included in-hospital death and fluid balance from 36 h to 84 h after ICU admission, given the average ICU length of stay is 3.3 days or 79 h [24]. For the fluid balance outcome, we excluded patients who died within the 84 h period.

In addition to our variables of interest, we identified potential confounders that would account for a patients’ physical characteristics, such as age and sex. We accounted to disease severity and comorbidity burden with the Acute Physiology and Chronic Health Evaluation (APACHE IV) score and the Charlson Comorbidity Index (CCI) in eICU, the Oxford Acute Severity of Illness Score (OASIS) and the Elixhauser Comorbidity Score in MIMIC III, APACHE II in AmsterdamUMC, and APACHE II in SNUH. Comorbidity information was not available for the AmsterdamUMC and SNUH databases.

### Data Analysis

After developing our VLI, we trained a Generalized Additive Model (GAM) to determine the relationship between VLI and in-hospital death and 36h to 84h fluid balance. We were interested in the nonlinear relationship between VLI and the two outcomes. Because of this, a spline term was introduced for VLI. In addition, we created quartiles of VLI as the explanatory variable.

In our models, we controlled for covariates, specifically severity of illness, chronic comorbidity (if available), age, and biological sex. The relationship between VLI and the outcome variables as determined by the GAM is visualized across all values of VLI and the statistical significance of the relationship is determined by the ANOVA test.

All analyses were done in R 3.6.1. Code for our research can be found on our Github repository [23].

## RESULTS

### Population Description

In Table 1, we show how we derived our final patient cohort from our inclusion and exclusion criteria.

**Table 1.** Study Population Selection for all four databases

| Dataset<br>(Number of<br>Patient<br>Encounters) | Inclusion<br>Criteria:<br>Sepsis<br>Patients | Exclusion criteria |  | In-Hospital<br>Death<br>Population | Fluid Balance Exclusion<br>Criteria |  | 36h-84h<br>Fluid<br>Balance<br>Population |
| --- | --- | --- | --- | --- | --- | --- | --- |
| eICU (200,859<br>patient encounters) | 41,071 | Diagnosis of bleeding or<br>receiving blood products | 2,825 | 3,246 | Missing 36h-84h Fluid<br>Balance<br>Patient<br>Mortality<br>before 84h | 957<br>83 | 2,206 |
|  |  | Excess Fluid Expulsion | 3,616 |  |  |  |  |
|  |  | RRT Patients | 840 |  |  |  |  |
|  |  | Patients Under 16 and<br>Erroneous Demographic<br>Data | 6 |  |  |  |  |
|  |  | Unreliable Fluid Data | 9,186 |  |  |  |  |
|  |  | Missing VLI Equation-<br>related data | 21,352 |  |  |  |  |
| MIMIC III<br>(n=53,423<br>patient<br>encounters) | 18,764 | Diagnosis of bleeding or<br>receiving blood products | 2,690 | 4,056 | Missing 36h-84h Fluid<br>Balance<br>Patient<br>Mortality<br>before 84h | 391<br>144 | 3,521 |
|  |  | Excess Fluid Expulsion | 251 |  |  |  |  |
|  |  | RRT Patients | 1,777 |  |  |  |  |
|  |  | Patients Under 16 and<br>Erroneous Demographic<br>Data | 820 |  |  |  |  |
|  |  | Missing VLI Equation-<br>related data | 9,170 |  |  |  |  |
| AmsterdamUMC<br>(n=23,172<br>patient<br>encounters) | 4,363 | Diagnosis of bleeding or<br>receiving blood products | 1,295 | 1,617 | Missing 36h-84h Fluid<br>Balance<br>Patient<br>Mortality<br>before 84h | 237<br>83 | 1,297 |
|  |  | RRT Patients | 318 |  |  |  |  |
|  |  | Patients Under 16 and<br>Erroneous Demographic<br>Data | 0 |  |  |  |  |
|  |  | Missing VLI Equation-<br>related data | 1,133 |  |  |  |  |
| SNUH<br>(n=16,082<br>patient<br>encounters) | 794 | RRT Patients | 541 | 146 | Missing 36h-84h Fluid<br>Balance<br>Patient<br>Mortality<br>before 84h | 0<br>17 | 129 |
|  |  | Patients Under 16 and<br>Erroneous Demographic<br>Data | 6 |  |  |  |  |
|  |  | Missing VLI Equation-<br>related data | 101 |  |  |  |  |

**Table 2.** Baseline Characteristics for the 36-84h fluid balance patient cohort in all four databases

| eICU |  | MIMIC III |  | Amsterdam |  | SNUH |  |
| --- | --- | --- | --- | --- | --- | --- | --- |
| n | 2206 | n | 3521 | n | 1297 | n | 129 |
| Age (mean (SD)) | 65.83<br>(16.09) | Age (mean (SD)) | 65.65<br>(16.69) | Age (%) |  | Age (mean (SD)) | 63.27<br>(15.69) |
| Gender = Male (%) | 1133<br>(51.4) | Gender = Male (%) | 1897<br>(53.9) | 18-39 | 166 ( 12.8) | Gender = Male (%) | 67 (51.9) |
| ICU Type (%) |  | ICU Type (%) |  | 40-49 | 158 ( 12.2) | ICU Type (%) |  |
| Cardiac ICU | 132 ( 6.0) | Cardiac ICU (CICU) | 426 (12.1) | 50-59 | 216 ( 16.7) | Thoracic Surgery ICU<br>(RICU) | 8 ( 6.2) |
| Cardiac ICU-Cardiothoracic ICU<br>(CCU-CTICU) | 145 ( 6.6) |  |  | 60-69 | 302 ( 23.3) |  |  |
| Cardiac Surgery ICU<br>(CSICU) | 80 ( 3.6) | Cardiac Surgery<br>Recovery Unit (CSRU) | 450 (12.8) | 70-79 | 286 ( 22.1) |  |  |
| Cardiothoracic ICU<br>(CTICU) | 14 ( 0.6) |  |  | 80+ | 169 ( 13.0) |  |  |
| Medicine ICU (MICU) | 400 (18.1) | Medicine ICU (MICU) | 1629<br>(46.3) | Gender = Male (%) | 768 ( 59.2) | Medicine ICU (MICU) | 81 (62.8) |
| Neuro ICU | 20 ( 0.9) |  |  | ICU Type = mixed<br>surgical-medical (%) | 1297<br>(100.0) |  |  |
| Med-Surg ICU | 1288<br>(58.4) |  |  |  |  |  |  |
| Surgery ICU (SICU) | 127 ( 5.8) | Surgery ICU (SICU) | 548 (15.6) |  |  | Surgical ICU (SICU) | 40 (31.0) |
|  |  | Trauma Surgical ICU<br>(TSICU) | 468 (13.3) |  |  |  |  |
| Weight (mean (SD)) | 81.07<br>(26.72) | Weight (mean (SD)) | 87.34<br>(25.08) | Weight (mean (SD)) | 77.75<br>(27.58) | Weight (mean (SD)) | 59.62<br>(11.75) |
| Height (mean (SD)) | 168.37<br>(11.26) | Height (mean (SD)) | 168.56<br>(10.66) | Height (mean (SD)) | 173.28<br>(10.99) | Height (mean (SD)) | 162.49<br>(8.36) |
| Body Surface Area<br>(mean (SD)) | 1.93 (0.33) | Body Surface Area<br>(mean (SD)) | 2.00<br>(0.31) | Body Surface Area<br>(mean (SD)) | 1.92 (0.29) | Body Surface Area<br>(mean (SD)) | 1.63<br>(0.19) |
| Vascular Leak Index<br>(median [IQR]) | -2.79 [-<br>5.24, -<br>1.46] | Vascular Leak Index<br>(median [IQR]) | -1.22 [-<br>2.35, -<br>0.35] | Vascular Leak Index<br>(median [IQR]) | -1.94 [-<br>3.48, -0.79] | Vascular Leak Index<br>(median [IQR]) | -0.80 [-<br>5.21,<br>1.13] |
| Apache IV Score (mean<br>(SD)) | 73.74<br>(25.93) | Oasis Score (mean<br>(SD)) | 36.55<br>(8.29) | Apache II Score (mean<br>(SD)) | 21.18<br>(6.61) | Apache II Score (mean<br>(SD)) | 26.16<br>(8.90) |
| Charlson Comorbidity<br>Index (mean (SD)) | 4.25 (2.88) | Elixhauser Comorbidity<br>Score (mean (SD)) | 10.60<br>(8.26) |  |  |  |  |
| First Hematocrit 18 hrs.<br>(mean (SD)) | 35.82<br>(6.82) | First Hematocrit 18 hrs.<br>(mean (SD)) | 33.44<br>(6.58) | First Hematocrit 18 hrs.<br>(mean (SD)) | 37 (7) | First Hematocrit 18 hrs.<br>(mean (SD)) | 30.43<br>(7.24) |
| Average Hematocrit 18-36 hrs. (mean (SD)) | 31.08<br>(5.49) | Average Hematocrit 18-36 hrs. (mean (SD)) | 30.08<br>(4.32) | Average Hematocrit 18-36 hrs. (mean (SD)) | 33 (6) | Average Hematocrit<br>18-36 hrs. (mean (SD)) | 29.23<br>(4.86) |
| Hospital Mortality (%) | 293 (13.3) | Hospital Mortality (%) | 529 (15.0) | Hospital Mortality (%) | 155 ( 12.0) | Hospital Mortality (%) | 24 (18.6) |
| Total Fluid Balance First<br>36 hrs. (mean (SD)) | 3302.50<br>(3977.82) | Total Fluid Balance First<br>36 hrs. (mean (SD)) | 7049.42<br>(6213.95) | Total Fluid Balance First<br>36 hrs. (mean (SD)) | 4008.54<br>(2625.10) | Total Fluid Balance<br>First 36 hrs. (mean<br>(SD)) | 1524.30<br>(1440.73) |
| Total Fluid Balance 36-84 hrs. (mean (SD)) | 768.82<br>(2676.75) | Total Fluid Balance 36-84 hrs. (mean (SD)) | 1375.12<br>(3647.28) | Total Fluid Balance 36-84 hrs. (mean (SD)) | 1973.01<br>(2626.22) | Total Fluid Balance 36-84 hrs. (mean (SD)) | 1026.19<br>(2192.98) |

The demographics, clinical characteristics, and outcomes for the patients in the four databases are presented in table 1 and supplementary table 1 for the fluid balance and in-hospital death outcomes, respectively.

The final distribution of VLI for each database for the in-hospital death cohort is presented in Supplementary Figure 1. VLI is left skewed with a small number of patients having positive leak indices. A positive leak index indicates that a patient’s hematocrit increased over time despite the administration of fluids.

### Association between VLI and in-hospital death

Using our GAM and analyzing VLI as quartiles, our results indicate that increasing VLI is associated with increased risk of in-hospital death in a dose-dependent manner. Patients in VLI Q4 had approximately 2.31 [CI: 1.71-3.12], 1.61 [CI: 1.26-2.05], and 2.13 [CI: 1.42-3.20] increased odds of dying in the hospital compared to patients in VLI Q1 for eICU, MIMIC, and Amsterdam, respectively. VLI Q2 and Q3 were also associated with increased odds of dying (Supplementary Fig 2).

Using our GAM and treating VLI as a continuous variable, we observe that in-hospital death changes with different values of VLI (Fig 1). There is high variability in extreme low values of VLI. However, we observe relatively low variability in VLI from −7 to 2 in eICU. In this range, there is an increase in hospital death from approximately 10% to 25%. In MIMIC, from a VLI of - 4 to 1, there is an increase in hospital death from approximately 14% to 23%. In Amsterdam, from a VLI of −3 to 1, there is an increase in-hospital death from approximately 13% to 25%. Overall, changes in the smoothed VLI are significantly associated with changes in in-hospital death in eICU (p<0.001), MIMIC (p=0.004), and Amsterdam (p=0.002).

**Figure 1.**
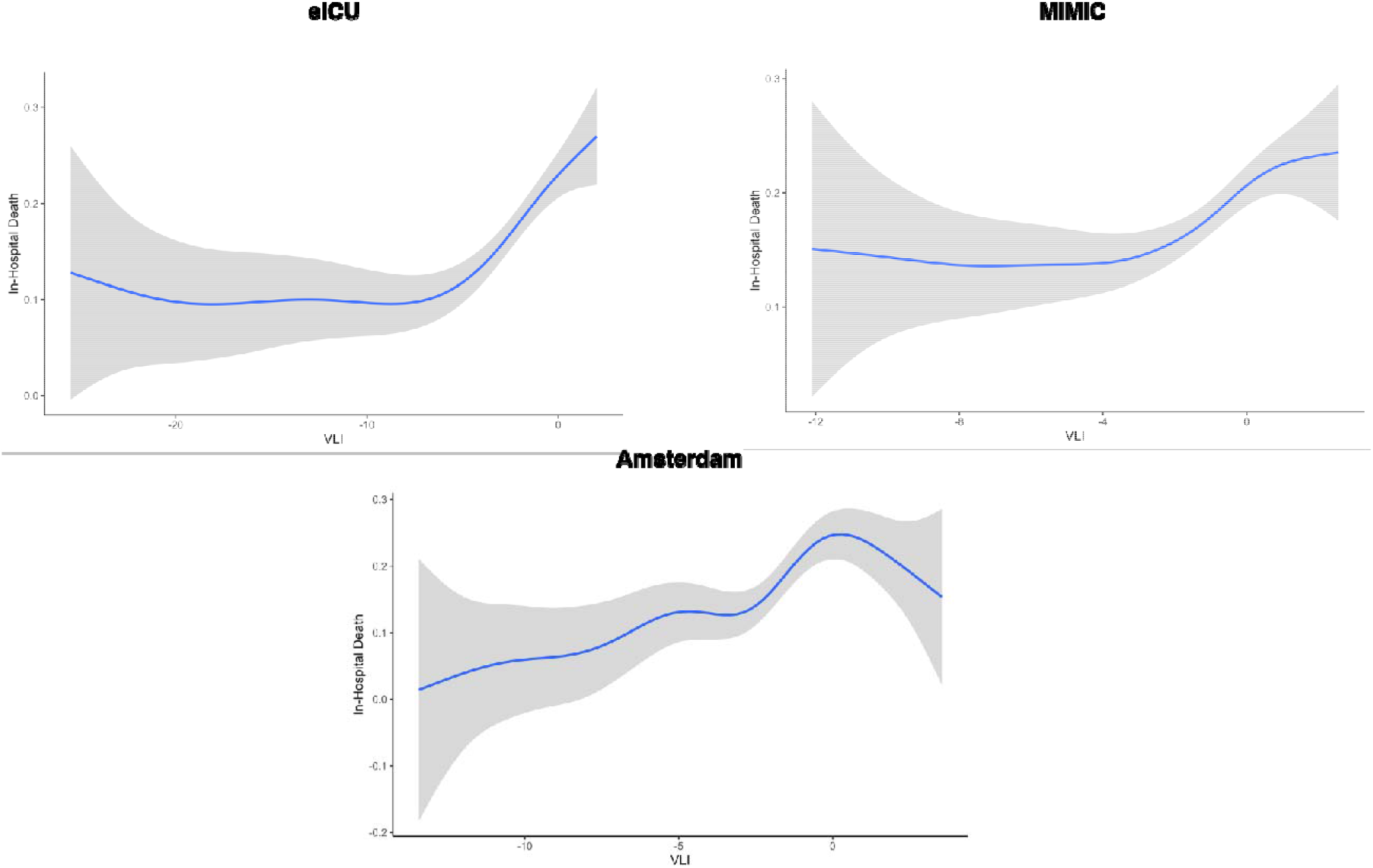
GAM fit for the association between VLI and proportion in in-hospital death for eICU, MIMIC, and Amsterdam. The blue line represents the mean proportion of in-hospital death while the gray shading is the 95% confidence interval.

See Supplementary Material for SNUH results.

### Association between VLI and 36h-84h Fluid Balance

Using our GAM and treating VLI as a continuous variable, we observe that 36h-84h fluid balance changes with different values of VLI (Fig 2). Again, there is high variability in extreme low values of VLI. From a VLI of −15 to a VLI of 2 in eICU, there is an increase in 36h-84 h fluid balance from approximately 0ml to 1500ml. In MIMIC, from a VLI of −4 to 1, there is an increase in 36h-84h fluid balance from approximately 700ml to 1600ml. In Amsterdam, from a VLI of −4 to 1, there is an increase in 36h-84h fluid balance from approximately 1500ml to 2200ml. Overall, the smoothed VLI is significantly associated with changes in 36h-84h fluid balance in eICU (p<0.001), MIMIC (p<0.001), Amsterdam (p = 0.043).

**Figure 2.**
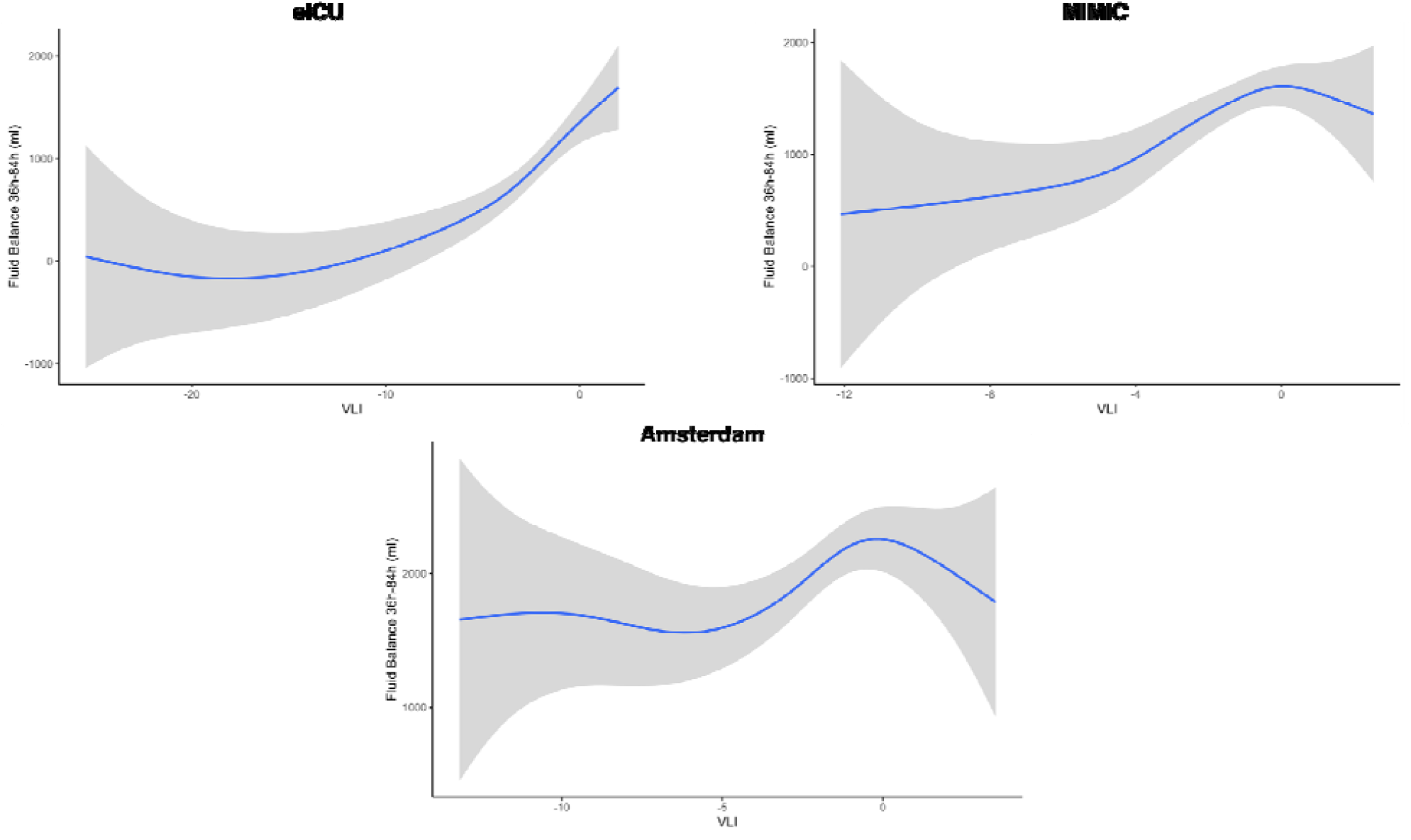
GAM fit for the association between VLI and fluid balance 36h-84h for eICU, MIMIC, and Amsterdam. The blue line represents the mean proportion of in-hospital death while the gray shading is the 95% confidence interval.

Treating VLI as quartiles, we observe that patients in VLI Q4 had approximately 1131ml (± 158ml, p<0.001), 685ml (± 180ml, p<0.001), and 528ml (± 206ml, p=0.011) increased 36h-84h fluid balance compared to patients in VLI Q1 for eICU, MIMIC, and Amsterdam respectively. Patients in VLI Q3 had approximately 800ml (± 158ml, p<0.001), 769ml (± 172ml, p<0.001), and 572ml (± 206ml, p=0.006) increased 36h-84h fluid balance compared to patients in VLI Q1 for eICU, MIMIC, and Amsterdam respectively.

See Supplementary Material for SNUH results.

## DISCUSSION

### Key findings

We developed an equation to identify patients with increased vascular leak and found that a higher VLI was associated with a higher fluid balance between 36h-84h of ICU care and a higher risk of in-hospital death in patients with negative VLI. The results for the patients with positive VLI varied across the databases due to considerations that will be discussed in the limitations section. Our analyses were replicated in several datasets from North America, Europe, and Asia, suggesting that the relationships observed are robust.

Our results suggest a causal relationship between VLI and risk for death according to updated Bradford Hill rules [25]. We found a dose-response relationship between the quartiles of VLI and risk for death. Our observations are consistent across several databases from around the world, and they fit well with current models of biological plausibility with respect to the severity of vascular leak, organ dysfunction, fluid administration, and risk for death. Whether increased VLI is a marker or mediator of increased mortality still needs exploration. We believe it is likely to be both.

### Clinical implications

Our findings suggest that clinicians may be able to determine whether a patient has high vascular leak within the first 36 hours of ICU care and thus who may be less likely to benefit from or could even suffer harm from additional IV fluid. Knowing a patient’s VLI might guide treatment, as we might restrict further IV fluids in the setting of high vascular leak, as extravasation of fluid and a positive fluid balance are associated with impaired organ function and a higher risk of death [26].

Recent studies suggest that fluid-sparing sepsis resuscitation may lead to improved outcomes. The CENSER trial compared the early use of norepinephrine to fluid-based sepsis resuscitation, finding that shock control, fluid balance, and risk of cardiogenic pulmonary edema were lower with a fluid restrictive approach [27]. Richard et al. prospectively randomized patients to receive sepsis resuscitation guided by invasive measures of fluid responsiveness, finding that limiting fluids to patients who were responsive was associated with a lower fluid balance, a lower risk for death, and more ventilator-free days, though the latter two differences were not statistically significant [28]. The FRESH randomized controlled trial compared standard fluid resuscitation with fluids guided by fluid responsiveness using a non-invasive monitor, finding that restricting IV fluids to patients who demonstrated fluid response was associated with lower fluid balance, lower risk for new renal replacement therapy, and lower risk for mechanical ventilation [29]. Our findings are consistent with and extend these observations, suggesting that patients with a higher VLI represent a population in whom IV fluids may be harmful, as they contribute to extravascular fluid accumulation. Our VLI does not require specialized equipment, which may make it attractive to care settings with limited resources.

A central goal of sepsis resuscitation is to restore or maintain blood flow to assure oxygen and metabolic substrate delivery to target tissues, supporting normal cellular function; this requires maintenance or restoration of the effective coordinated function of the macrocirculation, the microcirculation, and cellular function [30]. Increasing perfusion by augmenting cardiac output or arterial tone are crucial therapies. The fundamental goal of IV fluid is to increase venous return with the hope of increasing left ventricular end-diastolic volume, left ventricular stroke volume, cardiac output, and improvement of microcirculatory perfusion. However, a large proportion of critically ill patients do not demonstrate increases in cardiac output when they receive IV fluids [31].

Even in patients who demonstrate fluid responsiveness, IV fluids have only a small effect on blood volume and this effect may not be durable [32], [33]. Furthermore, even when IV fluids increase cardiac output, the increase may be offset by changes in other hemodynamic parameters, increasing oxygen delivery only slightly [34]. Because reduced endothelial barrier function and glycocalyx damage are characteristic of sepsis, a substantial proportion of infused fluids leaves the intravascular space within a short time [35], [36].

In situations like this, the harms of fluid may outweigh its benefits. Despite decades of research into identifying patients who are “fluid responsive,” clinicians still lack predictive tools that help identify patients who will benefit from IV fluids. The VLI we propose identifies patients who have increased risk of harm from IV fluids. We speculate that using VLI clinically may reduce harm associated with IV fluids. Further prospective studies are required to test this hypothesis formally.

### Limitations

Limitations common to studies that use electronic medical record databases include mistakes in charting data that must be input manually. Errors in collecting fluid data are especially prevalent given the amount of different fluid inputs that a patient receives and the difficulty of accurately measuring urine output. In eICU, we were able to limit our final patient cohort to those whose fluid intake data was reliable. However, this was not explicitly possible in other databases.

In addition, we found that most patients have a negative VLI. That is, the hematocrit decreased over time as they received IV fluids. In some patients, however, we observed an increase in hematocrit over the study period, even as the patients received IV fluids. Because we sought to exclude patients who received blood component transfusions, the reasons for the observed increase in hematocrit are uncertain. Due to the nature of our equation, patients with a rising hematocrit have the highest VLI and are classified in VLI Q4. The biological plausibility of placing these patients in the highest risk group is unclear. We believe that including patients with a rising hematocrit is one reason why we observe a high variability between VLI and the two outcomes of interest at high VLI values and why, in the AmsterdamUMC cohort, we observe decreasing mortality and decreasing 36h-84h fluid balance at the higher range of VLI.

Also, defining sepsis is particularly difficult, especially in a retrospective study when the diagnosis of sepsis may not be certain. In eICU, AmsterdamUMC, and MIMIC, we use relatively inclusive definitions while in SNUH, we were restricted to identifying sepsis in patients who received that diagnosis at admission.

A further limitation is the nature of our intuitive equation. We reasoned from physiological principles and attempted to normalize by using the body surface area as a surrogate for circulating blood volume. We recognize that body surface area may be an unreliable surrogate [37]. Nevertheless, we sought an index that uses easily available data in order to facilitate bedside use, especially in low- or middle-income settings where more sophisticated monitoring may not be available. Future work should validate our VLI prospectively.

In the different hospitals that represent these four databases, fluid administration practices were quite different. In addition, depending on fluid administration practices, 36h to 84h fluid balance and death can be significantly dependent on fluid balance in the first 36h. This makes it difficult to cleanly describe the relationship between VLI and 36h to 84h fluid balance.

Finally, while the patient population size was sufficiently large in the eICU, MIMIC, and AmsterdamUMC databases, the SNUH database had a very small final cohort size. The relationships seen in the other three datasets were generally maintained in the SNUH dataset, but the low sample size led to high variability in estimates.

## Conclusions

Using a VLI derived from changes in hematocrit and net fluid balance within the first 36 hours of a patient’s ICU care, we were able to estimate vascular leak and identify a population with a higher risk for dying and increased 36h-84h fluid balance in the hospital using a GAM that controlled for disease severity and chronic comorbidities. Future studies should validate our VLI and test whether using our VLI to guide therapy may result in patient-centered benefit. Other future studies should examine other outcomes, such as hospital length of stay, risk for new renal replacement therapy or mechanical ventilation, and others.

## Data Availability

Code can be found at https://github.com/theonesp/vol_leak_index

## Acknowledgements

The project was conceived, designed and conducted during the 2019 fall course HST.953 Collaborative Data Science in Medicine at the Harvard-MIT Division of Health Science and Technology. Yueh-Hsu, Runyu Hong, Shari B. Brosnahan, John Munger, David A. Kaufman, and Kimiko Huang supported this research during the 2019 NYU Health Datathon. LAC is funded by the National Institute of Health through R01 EB017205.

