## Supplemental Material for "A novel Vascular Leak Index identifies sepsis patients with a higher risk for in-hospital death and fluid accumulation"

**Information for Each Database and IRB Approval/Exemption**

The eICU database includes 200,859 patient encounters for 139,367 unique patients admitted to the ICU in 2014 and 2015 from across the U.S. (14). 208 hospitals contributed data, and the median age of patients was 65 years old. The population included similar numbers of men and women (male=108,8879, 53.96%), and ethnicity was heavily distributed to Caucasian (n=155,285, 77.31%) and African American descent (n=21,308, 10.61%). Patients were mainly admitted to medical and surgical ICUs (n=113,222, 56.37%). Sepsis was the most common of the APACHE diagnoses (n=18,087, 16.40%), double that of the next most common diagnoses including cerebrovascular accident, cardiac arrest, and acute coronary syndrome.

The study was exempt from institutional review board approval due to the retrospective design, lack of direct patient intervention, and the security schema, for which the re-identification risk was certified as meeting safe harbor standards by an independent privacy expert (Privacert, Cambridge, MA) (Health Insurance Portability and Accountability Act Certification no. 1031219-2).

The MIMIC-III database includes about 60,000 patients admitted to critical care units in the Beth Israel-Deaconess Medical Center from 2001 to 2012. The database includes information for adult patients, aged 16-years-old or older, admitted between 2001 and 2012, as well as for neonates admitted between 2001 and 2008. From the data on adult patients, median age across the critical care units was 65.8 years, and the primary codes for adult admissions were for coronary atherosclerosis of native coronary artery (7.1%), unspecified septicemia (4.2%), and subendocardial infarction (3.6%).

The data in MIMIC-III and MIMIC-CXR have been de-identified, and the institutional review boards of the Massachusetts Institute of Technology (No. 0403000206) and Beth Israel Deaconess Medical Center (2001-P-001699/14) both approved the use of the database for research.

We used version 1.0 of the AmsterdamUMCdb database in this study. It includes 23,172 intensive care unit admissions of 20,109 adult patients from 2003-2016 in the Amsterdam University Medical Center. The dataset contained 64% men (and a median age of approximately 63 years old. Around 12% of admissions were sepsis patients, which is the second most common diagnosis behind cardiothoracic surgery (26%).

The Medical Research Ethics Committee of VU university medical center determined that the AmsterdamUMCdb study was exempt from their review and was not subject to the Dutch Medical Research Involving Human Subjects Act (WMO). The process of developing AmsterdamUMCdb was audited by an external team led by a member of the privacy expert group at the Netherlands Federation of UMCs. The Ethics in Intensive Care Medicine group provided external ethics review and appraisal. The use of AmsterdamUMCdb is exempt from institutional review board approval due to a combination of de-identification, contractual and governance strategies where re-identification is not reasonably likely and can therefore be considered as anonymous information in the context of the General Data Protection Regulation (GDPR).

The SNUH dataset contains 16,082 ICU admissions between 2017 and 2020 in the Seoul National University Hospital. The dataset contains 58% men and a median age of approximately 63 years old. Only about 5% of all admissions were explicitly diagnosed with sepsis.

The data in the SNUH dataset have been de-identified, and the institutional review boards of the Seoul National University Hospital have approved the use of this data for our research (SNUH 2106-118-1228).

| eICU | | MIMIC III | | Amsterdam | | SNUH | |
|---|---|---|---|---|---|---|---|
| n | 3246 | n | 4056 | n | 1617 | n | 146 |
| Age (mean (SD)) | 66.00 (16.48) | Age (mean (SD)) | 65.85 (16.67) | Age (%) | | Age (mean (SD)) | 63.37 (15.78) |
| Gender = Male (%) | 1643 (50.6) | Gender = Male (%) | 2170 (53.5) | 18-39 | 198 ( 12.2) | Gender = Male (%) | 76 (52.1) |
| ICU Type (%) | | ICU Type (%) | | 40-49 | 189 ( 11.7) | ICU Type (%) | |
| Cardiac ICU | 179 ( 5.5) | Cardiac ICU (CICU) | 498 (12.3) | 50-59 | 258 ( 16.0) | Thoracic Surgery ICU (RICU) | 10 ( 6.8) |
| Cardiac ICU-Cardiothoracic ICU (CCU-CTICU) | 285 ( 8.8) | | | 60-69 | 367 ( 22.7) | | |
| Cardiac Surgery ICU (CSICU) | 123 ( 3.8) | Cardiac Surgery Recovery Unit (CSRU) | 450 (12.8) | 70-79 | 366 ( 22.6) | | |
| Cardiothoracic ICU (CTICU) | 23 ( 0.7) | | | 80+ | 239 ( 14.8) | | |
| Medicine ICU (MICU) | 676 (20.8) | Medicine ICU (MICU) | 1629 (46.3) | Gender = Male (%) | 945 ( 58.4) | Medicine ICU (MICU) | 90 (61.6) |
| Neuro ICU | 28 ( 0.9) | | | ICU Type = mixed surgical-medical (%) | 1617 (100.0) | | |
| Med-Surg ICU | 1775 (54.7) | | | | | | |
| Surgery ICU (SICU) | 157 ( 4.8) | Surgery ICU (SICU) | 599 (14.8) | | | Surgical ICU (SICU) | 46 (31.5) |
| | | Trauma Surgical ICU (TSICU) | 488 (12.0) | | | | |
| Weight (mean (SD)) | 81.26 (26.94) | Weight (mean (SD)) | 86.38 (24.97) | Weight (mean (SD)) | 77.17 (25.87) | Weight (mean (SD)) | 60.09 (12.61) |
| Height (mean (SD)) | 168.22 (11.48) | Height (mean (SD)) | 168.48 (10.78) | Height (mean (SD)) | 172.94 (11.47) | Height (mean (SD)) | 162.60 (8.59) |
| Body Surface Area (mean (SD)) | 1.93 (0.33) | Body Surface Area (mean (SD)) | 1.99 (0.31) | Body Surface Area (mean (SD)) | 1.91 (0.28) | Body Surface Area (mean (SD)) | 1.64 (0.20) |
| Vascular Leak Index (median [IQR]) | -2.79 [-5.34, -1.45] | Vascular Leak Index (median [IQR]) | -1.22 [-2.43, -0.34] | Vascular Leak Index (median [IQR]) | -1.94 [-3.53, -0.79] | Vascular Leak Index (median [IQR]) | -0.80 [-4.95, 1.65] |
| Apache IV Score (mean (SD)) | 72.75 (26.84) | Oasis Score (mean (SD)) | 36.17 (8.52) | Apache II Score (mean (SD)) | 21.07 (6.83) | Apache II Score (mean (SD)) | 26.98 (9.05) |
| Charlson Comorbidity Index (mean (SD)) | 4.28 (2.88) | Elixhauser Comorbidity Score (mean (SD)) | 10.59 (8.32) | | | | |

| | | | | | | | |
|---|---|---|---|---|---|---|---|
| First Hematocrit 18 hrs. (mean (SD)) | 35.72 (6.81) | First Hematocrit 18 hrs. (mean (SD)) | 33.36 (6.59) | First Hematocrit 18 hrs. (mean (SD)) | 37 (7) | First Hematocrit 18 hrs. (mean (SD)) | 29.95 (7.04) |
| Average Hematocrit 18-36 hrs. (mean (SD)) | 31.17 (5.45) | Average Hematocrit 18-36 hrs. (mean (SD)) | 30.11 (4.39) | Average Hematocrit 18-36 hrs. (mean (SD)) | 33 (6) | Average Hematocrit 18-36 hrs. (mean (SD)) | 29.12 (4.72) |
| Hospital Mortality (%) | 521 (16.1) | Hospital Mortality (%) | 713 (17.6) | Hospital Mortality (%) | 271 ( 16.8) | Hospital Mortality (%) | 39 (26.7) |
| Total Fluid Balance First 36 hrs. (mean (SD)) | 3165.10 (3559.74) | Total Fluid Balance First 36 hrs. (mean (SD)) | 6844.01 (6239.66) | Total Fluid Balance First 36 hrs. (mean (SD)) | 3887.88 (2676.60) | Total Fluid Balance First 36 hrs. (mean (SD)) | 1632.11 (1443.04) |

**Supplementary Table 1.** Baseline Characteristics for the in-hospital mortality patient cohort in all four databases

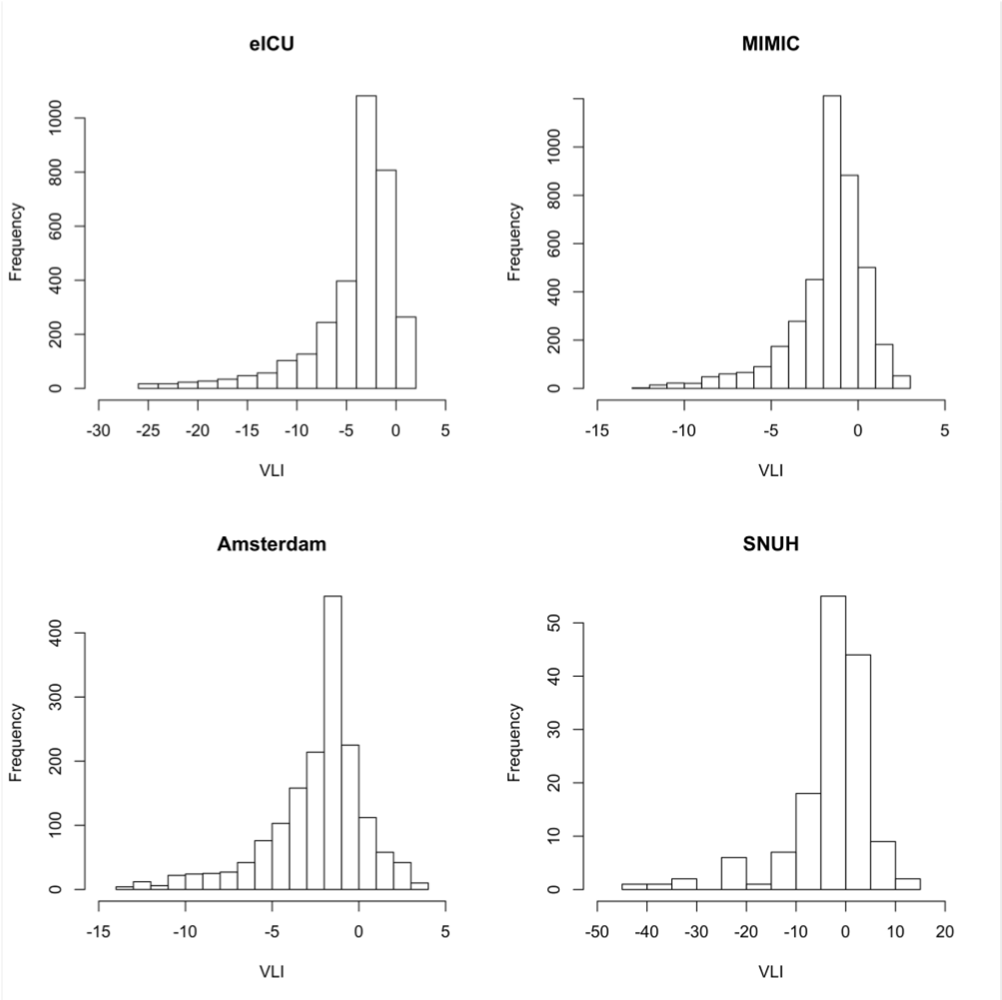


**Supplementary Figure 1**. Distribution of VLI for all four databases for the in-hospital mortality patient cohort.

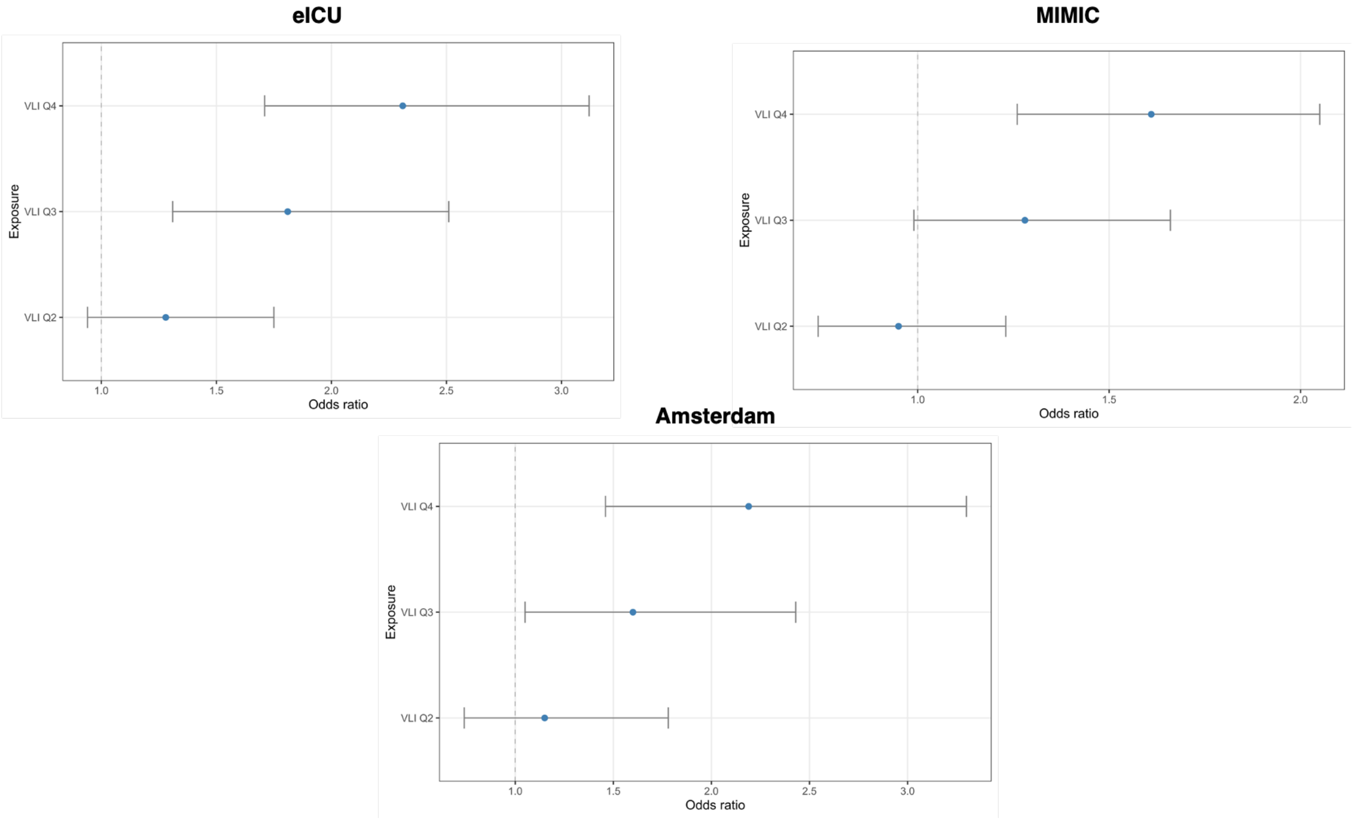


**Supplementary Figure 2**. Mortality Odds Ratio for VLI Quartile 2,3, and 4 in comparison to VLI Q1 for eICU, MIMC, and Amsterdam

### SNUH

#### Population description

Of the 16,082 patient encounters from the SNUH database, we studied a cohort of 129 patient and 146 patient encounters for the fluid balance outcome and in-hospital mortality outcome respectively. This cohort's demographics, clinical characteristics, and outcomes are summarized in Table 2.

#### Association between VLI and in-hospital mortality

Using our GAM and treating VLI as quartiles, our results indicate that increasing VLI is associated with increased risk of in-hospital mortality in a dose-dependent manner. Patients in VLI Q4 had approximately 2.28 [CI: 0.62-8.39] increased odds of dying in the hospital compared to patients in VLI Q1. Other VLI quartiles were associated with increased odds of dying, at approximately 2.26 [CI: 0.59-8.71] and 1.16 [CI: 0.47-5.94] increased odds for VLI Q3 and VLI Q2 compared to VLI Q1, respectively (Supplementary Figure 3).

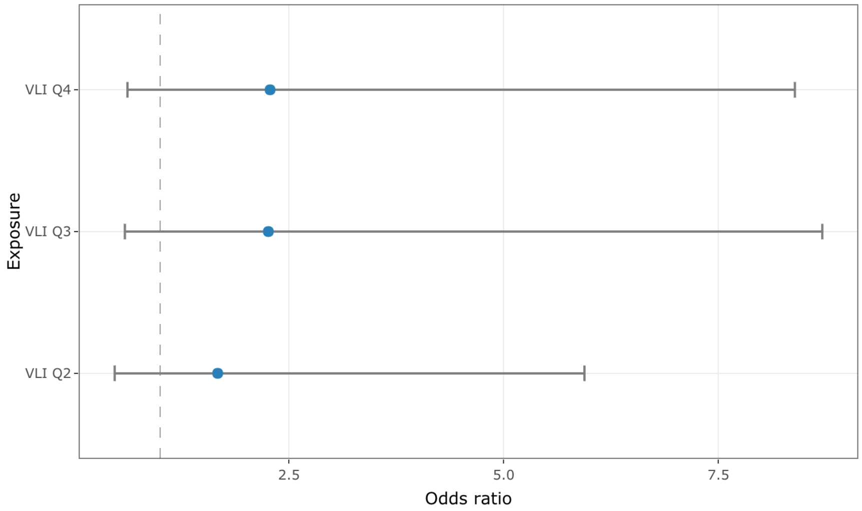


**Supplementary Figure 3.** Association between VLI and in-hospital mortality.

Using our GAM and treating VLI as a continuous quantity, we show how in-hospital mortality changes with different values of VLI (Supplementary Figure 4). In the region of lowest variability, from a VLI of -20 to a VLI of 10, there is an increase in in-hospital mortality from approximately 12% to 36%. Overall, the smoothed VLI is associated with the changes in-hospital death (p = 0.234).

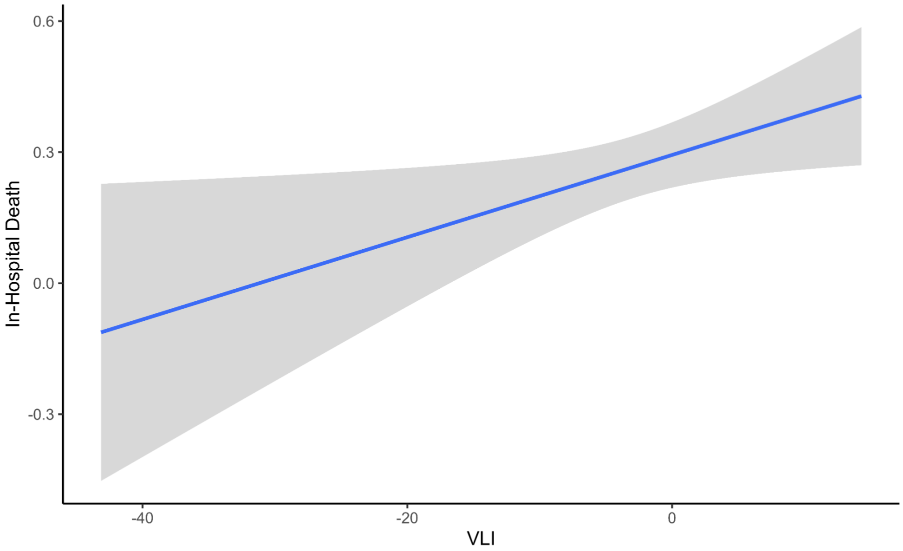


**Supplementary Figure 4.** Generalized additive model (GAM) for association between VLI and risk for in-hospital mortality. The blue line represents the mean proportion of in-hospital death while the gray shading is the 95% confidence interval.

**Association between VLI and 36h-84h Fluid Balance**

Using our GAM and treating VLI as a continuous quantity, we show how 36h-84h fluid balance changes with different values of VLI (Supplementary Figure 5). In the region of lowest variability, from a VLI of -10 to a VLI of 5, there is an increase in in-hospital mortality from approximately 300ml to 1000ml. Overall, the smoothed VLI is associated with the changes 36h-84h fluid balance (p = 0.338). The low sample size in the SNUH dataset yields results with high variability.

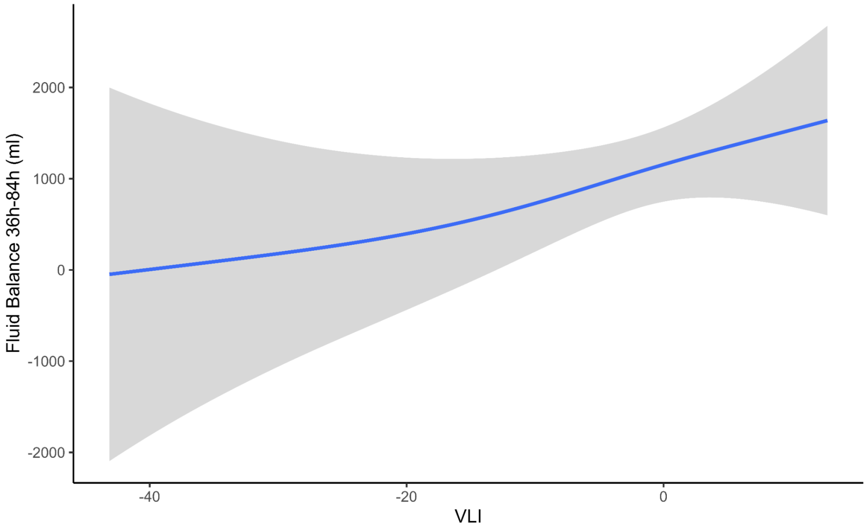


**Supplementary Figure 5.** Generalized additive model (GAM) for association between VLI and risk for 36h-84h Fluid Balance. The blue line represents the mean proportion of in-hospital death while the gray shading is the 95% confidence interval.

Treating VLI as quartiles, we show that being in VLI Q4, VLI Q3, and VLI Q2 increases 36h-84h fluid balance by 347ml (± 526ml, p=0.51), 1743ml (± 586ml, p<0.001), 830ml (± 483ml, p=0.089) respectively in comparison to VLI Q1.
